# Geospatial AI targeting of tuberculosis active case finding in Pune, India: a quasi-experimental evaluation

**DOI:** 10.64898/2026.11.11.26367599

**Authors:** Ujjwal Rao, Radhakishan Pawar, Archana Beri, Sarabjit Chadha, Himanshu Vashistha, Mrinalini Das

## Abstract

**BACKGROUND:** Active case finding (ACF) in India screens large populations to find few cases. AI-NETRA predicts neighbourhood-level tuberculosis (TB) risk from routine notification data and generates field microplans.

**OBJECTIVE:** To compare the yield and efficiency of AI-guided and standard ACF, and frontline acceptability.

**DESIGN:** Prospective quasi-experimental evaluation in four TB Units of Pune district, comparing two-week AI-guided sweeps (April to May 2026) with the routine campaign in the same units (December 2024 to January 2025). Primary endpoint: bacteriologically confirmed pulmonary TB per persons screened; secondary: all-forms yield, presumptive yield, referral precision, number needed to screen (NNS) and cost per case. Acceptability: 16 focus groups (n=100) and a survey of 101 ASHAs.

**RESULTS:** AI-guided teams screened 66,095 people and confirmed 10 pulmonary cases, against 9 among 177,322 in the standard campaign (rate ratio 2.98, 95% CI 1.09 to 8.29); NNS was 6,610 versus 19,702 and cost per case 66% lower. All-forms detection did not differ (15 versus 32; RR 1.26, 95% CI 0.63 to 2.39); the standard campaign had mobile X-ray. 96% of ASHAs reported finding the same presumptive cases while screening fewer people.

**CONCLUSION:** Geospatial targeting raised confirmed yield per person and lowered NNS; total detection did not differ. A concurrent, like-for-like trial is warranted.

## Introduction

India carries about a quarter of the global tuberculosis (TB) burden, and the National TB Elimination Programme (NTEP) has made active case finding (ACF) a central instrument for reaching people who do not present to care, most recently through the 100-day intensified campaign launched in December 2024.^[1-3]^ ACF is effective but inefficient when screening is spread evenly across a population: national analyses report a median number needed to screen (NNS) in the thousands, with yield below programme targets in most states.^[4,5]^ In Pune district, a routine campaign screens about half a million people to detect roughly 100 cases, an NNS near 5,000.^[6]^

TB is not evenly distributed. It clusters in identifiable neighbourhoods, and WHO screening guidance asks programmes to prioritise where transmission concentrates.^[7-9]^ The geospatial tools now used in Indian programmes resolve risk to the block or ward, which helps allocate effort across a district but offers no guidance inside a dense ward, where a whole-ward campaign is rarely feasible.^[10]^ AI-NETRA (Froncort.AI, Pune) addresses the gap below the ward: it scores TB risk for each cell of the H3 hexagonal grid from routine Nikshay notification data and assembles the highest-risk cells into a microplan that a frontline worker can follow.^[11]^ In retrospective validation against three Pune campaigns, its prioritised cells captured 89% of campaign-detected cases from under one-third of the screened population, and its taluka-level rank correlation with observed cases rose from 0.55 to 0.78 across successive campaigns.^[6]^ Whether that translates into yield in the field had not been tested.

We report a prospective quasi-experimental evaluation comparing AI-guided ACF with the standard campaign in four TB Units of Pune district, with a pre-specified bacteriological primary endpoint, and a mixed-methods assessment of acceptability and usability among the workers who executed the plans. The study corresponds to an early-stage live evaluation of an AI decision support system and is reported against TREND, with COREQ for the qualitative component.^[12-14]^

## Methods

### Design and setting

This was a prospective, quasi-experimental, controlled before-after comparison of two ACF planning strategies in the same geographies: AI-guided micro-targeting versus the standard blanket campaign. It contrasts planning strategies, not exposures, and is reported as a non-randomised evaluation. Four TB Units (TUs) of Pune district, Maharashtra, were selected to span the rural and urban ACF context and the range of predicted yield: Bhor and Shirur (rural) and Haveli PMC and Haveli PCMC (urban, within the Pune and Pimpri-Chinchwad municipal corporations).

### The planning tool

AI-NETRA represents the district as H3 hexagons and scores each hexagon’s TB risk from the Nikshay notification register. The version 1 model was trained on bacteriologically confirmed notifications from 2022 to 2025 geocoded to residential address; about 92% of records were geocoded, 16% to building precision and 72% to area precision, and all were retained. Features computed for each hexagon and month combine recent case history with temporal decay, case characteristics weighted towards pulmonary disease, spatial spillover from adjacent hexagons, cold-start inference from structural proxies where notification history is sparse, and programmatic vulnerability indicators; a two-stage gradient-boosted hurdle model, a calibrated probability that the hexagon will notify any case in the following month multiplied by a Poisson expected count, ranks hexagons for the month ahead. Hexagons in the top 10% of scores are classed high priority and those between the 10th and 30th percentiles medium priority. Navigation hotspots are listed at H3 resolution 10 (about 123 x 123 m equivalent) and the screening population, the NNS denominator, is computed at resolution 9 (about 325 x 325 m) for operational feasibility. Each microplan lists hotspots by landmark, address and map link, with the residential clusters they contain, cases notified in the previous six months, a predicted case range and the number of people to screen (Supplementary Appendix S1). A single locked model version generated the plans before the sweep. An open-source implementation of the model is available under the Apache-2.0 licence (Data and code availability). The tool decides where teams go; it does not change who is screened within a household or how presumptive TB is identified, sampled, tested or treated, which followed the NTEP algorithm.

### AI-guided arm

In each unit, ASHAs and TB staff trained before deployment conducted a two-week sweep of the microplan areas between April and May 2026 (rural units in April, urban units in May). All high-priority hexagons were covered comprehensively; about one-third of medium-priority hexagons were covered, selected by rank with geographic stratification; and non-hotspot areas adjacent to the plan served as an opportunistic specificity check. Teams applied the NTEP symptom screen house to house, and presumptive TB was referred for sputum collection and nucleic acid amplification testing (NAAT; Truenat or CBNAAT). The district’s mobile chest X-ray van was not deployed with the sweep; chest radiography was available only on referral to a health facility.

### Standard arm

The comparator was the routine district ACF campaign conducted in the same four units from December 2024 to January 2025 (14 days): house-to-house symptom screening of the whole target population, sputum microscopy for presumptive TB with NAAT for a subset, and mobile chest X-ray available. Unit-level counts of persons screened, presumptive TB identified and tested, and cases diagnosed by category were taken from the campaign’s TB-4 reporting (Supplementary Table S2).

### Outcomes

The pre-specified primary endpoint was bacteriologically confirmed pulmonary TB per persons screened, analysed on the as-screened population. In the AI arm this was NAAT-positive pulmonary TB, the NTEP standard definition, and every bacteriologically confirmed pulmonary case in that arm was NAAT-positive; in the standard arm it was the TB-4 reporting category of sputum smear- or NAAT-positive pulmonary TB, which also admits smear-only diagnoses and is therefore, if anything, the broader of the two. Secondary endpoints were all-forms TB (bacteriologically confirmed or clinically diagnosed, pulmonary or extrapulmonary), which served as the total-case-detection measure and as a conservative sensitivity analysis favouring the standard arm; presumptive yield; referral precision (confirmed pulmonary cases per presumptive tested); NNS; and cost per confirmed case, applying the programme’s ACF unit cost per person screened to each arm’s screening effort and yield (Supplementary Appendix S1). Cases in the AI arm were attributed to the TB Unit in which they were identified during the sweep.

### Sample size and statistical analysis

The field window fixed the achievable number of confirmed cases, so the study was powered for estimation rather than hypothesis testing: at 80% power, detecting a rate ratio of 1.25 would need about 640 confirmed cases, 1.5 about 200, 2.0 about 75 and 3.0 about 35, and an exact 95% interval around 10 events spans 4.8 to 18.4. Yields are reported per 10,000 screened with exact (Garwood) Poisson intervals, and NNS as persons screened per case with intervals obtained by inverting the Poisson interval.^[15]^ Rate ratios comparing arms used the exact conditional (binomial) form of the Poisson test with exact intervals; presumptive yield used Fisher’s exact test and log-binomial regression; rural and urban contrasts were pre-specified subgroups. Pre-specified sensitivity analyses were leave-one-out omission of each confirmed case and reconstruction of the urban denominators, which were population-covered counts rather than household-enumerated screens. Analyses used Python 3 (SciPy 1.11).

### Acceptability and usability

Three instruments were used. Sixteen focus group discussions (FGDs) with 100 frontline workers were held across the four units between 17 April and 11 May 2026, facilitated in Marathi by implementation-partner staff around a hands-on demonstration of a microplan. A moderator and a note-taker recorded responses on a structured template covering current ACF practice, the AI plan, eight mobile-usability statements rated from 1 to 5, field context, workflow and supervision, trust, a 17-item feature-priority vote and overall acceptability, with the group as the unit of analysis; reporting follows COREQ (Supplementary Appendix S4). A time-and-motion study timed 57 workers through the five steps of opening a plan and starting navigation to a hotspot. A structured post-campaign survey of 101 ASHAs, administered in Marathi as an online form between 3 and 30 June 2026 after field use, is treated as the primary in-practice acceptability signal. Quantitative and qualitative strands were integrated in a joint display following GRAMMS.^[16]^ A pre-deployment survey of programme leadership is reported separately.^[6]^

### Ethics and registration

The prospective phase was approved by the Ethics Committee of the Jehangir Clinical Development Centre, Pune (JCDC/BHR/26/005, 13 March 2026). Routine programme data were used under NTEP data-governance arrangements, and FGD and survey participants gave informed consent. The protocol and statistical analysis plan, including the primary endpoint, were fixed before the sweep. The study evaluated a planning method within routine programme operations and did not alter individual clinical management; it was not registered as a clinical trial.

## Results

### Screening cascade

Across the four units, AI-guided teams visited 17,817 households and screened 66,095 people, identifying 326 presumptive TB cases (0.49%; 1.83 per 100 households), of whom 325 (99.7%) were tested (Table 1). Fifteen TB cases of all forms were diagnosed (4.6% of those tested): 11 pulmonary, of which 10 were NAAT-positive (7 by Truenat, 3 by CBNAAT) and 1 was diagnosed on chest radiography, and 4 extrapulmonary (3 pleural, diagnosed radiologically, and 1 abdominal, on cytopathology). The 10 NAAT-positive pulmonary cases met the primary definition: Shirur 1, Bhor 0, Haveli PMC 7 and Haveli PCMC 2. Cases were aged 14 to 70 years (median 34), 9 were male, 10 were notified as residents of urban slums, and all 15 started treatment within three days of diagnosis (Supplementary Table S3b). The AI arm screened 37% as many people as the standard campaign had in the same units (177,322), and each sweep was completed within two weeks.

**Table 1.** Screening cascade and bacteriologically confirmed pulmonary TB yield by TB Unit, AI-guided arm.

| TB Unit | Setting | Households visited | Persons screened | Presumptive TB | Tested | Diagnosed, all forms | Confirmed pulmonary (primary) | Yield per 10,000 (95% CI) | NNS |
|---|---|---|---|---|---|---|---|---|---|
| Shirur | Rural | 855 | 3,710 | 31 | 31 | 2 | 1 | 2.70 (0.07 to 15.0) | 3,710 |
| Bhor | Rural | 815 | 3,098 | 23 | 23 | 0 | 0 | 0 (0 to 11.9) | >840* |
| Haveli PMC | Urban | 7,004 | 27,047 | 204 | 204 | 9 | 7 | 2.59 (1.04 to 5.33) | 3,864 |
| Haveli PCMC | Urban | 9,143 | 32,240 | 68 | 67 | 4 | 2 | 0.62 (0.08 to 2.24) | 16,120 |
| Total |  | 17,817 | 66,095 | 326 | 325 | 15 | 10 | 1.51 (0.73 to 2.78) | 6,610 (3,594 to 13,783) |

Exact (Garwood) Poisson intervals; NNS = persons screened per confirmed pulmonary case. \*Bhor recorded no case, so its NNS is a lower bound from the upper interval limit. Urban denominators are population-covered counts (Methods). Field data extract, 29 June 2026.

### Primary endpoint

The pooled yield of bacteriologically confirmed pulmonary TB was 1.51 per 10,000 screened (95% CI 0.73 to 2.78), an NNS of 6,610 (95% CI 3,594 to 13,783); unit-level yields were imprecise, as expected at these counts (Table 1). Against the standard campaign (9 cases among 177,322 screened; 0.51 per 10,000), the rate ratio was 2.98 (95% CI 1.09 to 8.29; exact p=0.019), and the NNS was 6,610 versus 19,702, 66% lower (Table 2; Figure 1). The advantage was carried by the urban units (9 of 59,287 versus 7 of 114,816; RR 2.49, 95% CI 0.83 to 7.87), was large but imprecise in Shirur (1 versus 1; RR 11.5) and absent in Bhor (0 versus 1). Omitting any single AI-arm case reduced the rate ratio to 2.68 (95% CI 0.94 to 7.63; p=0.058).

**Table 2.** AI-guided versus standard ACF in the same four TB Units: cases, rate ratios, NNS and cost.

| Outcome | AI-guided: cases / screened | Standard: cases / screened | Rate ratio (95% CI) | Exact p |
|---|---|---|---|---|
| Bacteriologically confirmed pulmonary TB (primary) |  |  |  |  |
| Bhor | 0 / 3,098 | 1 / 19,842 | 0 (0 to 250) | 1.00 |
| Shirur | 1 / 3,710 | 1 / 42,664 | 11.5 (0.15 to 903) | 0.15 |
| Haveli (PMC and PCMC) | 9 / 59,287 | 7 / 114,816 | 2.49 (0.83 to 7.87) | 0.069 |
| Pooled, four units | 10 / 66,095 | 9 / 177,322 | 2.98 (1.09 to 8.29) | 0.019 |
| Pooled, any one AI-arm case omitted | 9 / 66,095 | 9 / 177,322 | 2.68 (0.94 to 7.63) | 0.058 |
| All forms of TB (secondary) |  |  |  |  |
| Bhor | 0 / 3,098 | 2 / 19,842 | 0 (0 to 34.1) | 1.00 |
| Shirur | 2 / 3,710 | 4 / 42,664 | 5.75 (0.52 to 40.1) | 0.08 |
| Haveli (PMC and PCMC) | 13 / 59,287 | 26 / 114,816 | 0.97 (0.46 to 1.95) | 1.00 |
| Pooled, four units | 15 / 66,095 | 32 / 177,322 | 1.26 (0.63 to 2.39) | 0.51 |
| Efficiency (primary endpoint) |  |  |  |  |
| NNS (95% CI) | 6,610 (3,594 to 13,783) | 19,702 | 66% lower |  |
| Cost per confirmed case | Rs 9.3 lakh | Rs 27.6 lakh | 66% lower |  |

Standard arm: routine district ACF campaign, December 2024 to January 2025, same four units, from TB-4 reporting (Haveli PMC and PCMC reported together). Exact conditional binomial rate ratios and p-values. AI-arm primary cases are NAAT-positive pulmonary TB (every bacteriologically confirmed pulmonary case in the AI arm was NAAT-positive); standard-arm primary cases are the TB-4 category of sputum smear- or NAAT-positive pulmonary TB. Cost at the programme's ACF unit cost per person screened (Supplementary Appendix S1).

**Figure 1.**
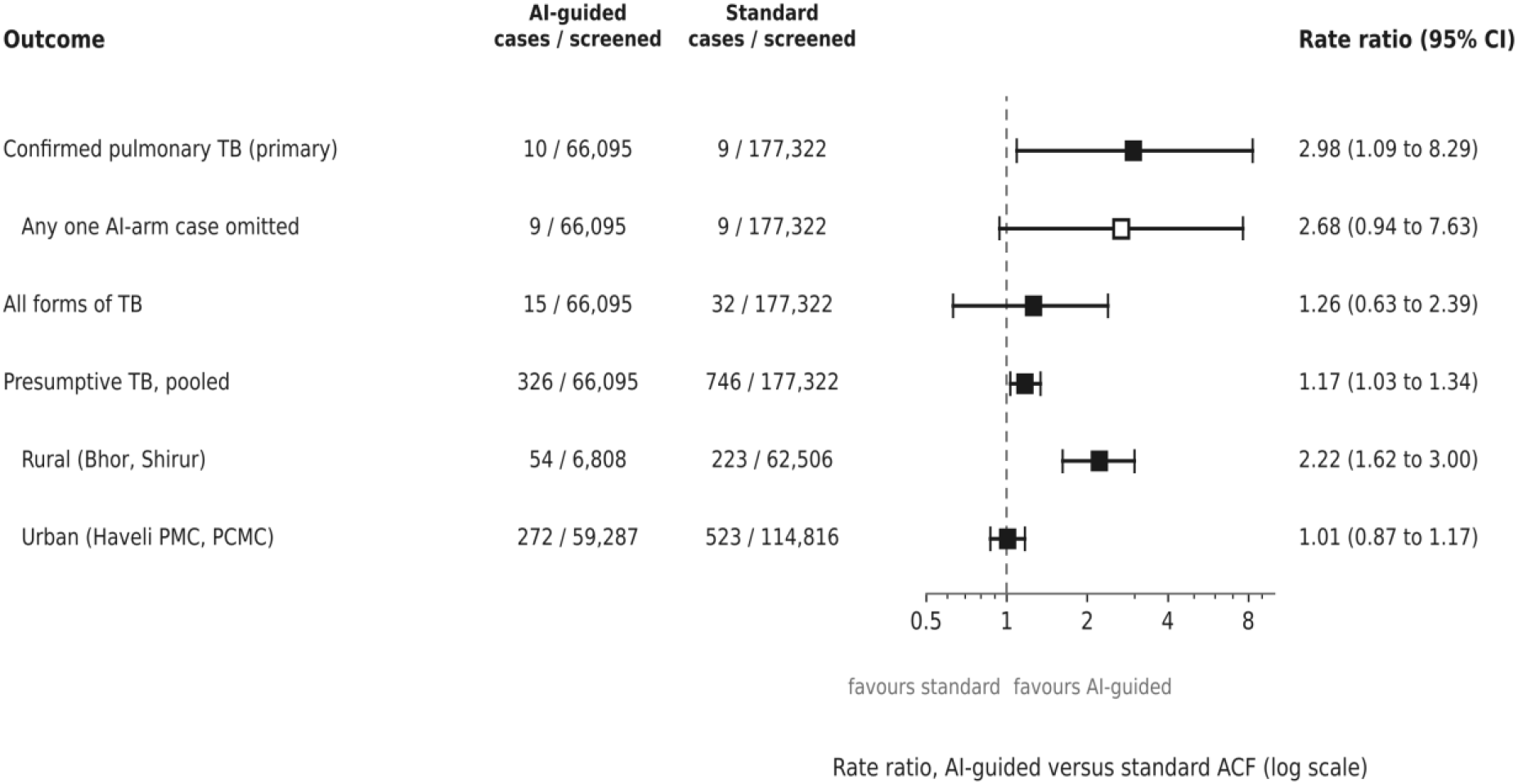
Rate ratios for AI-guided versus standard ACF, four TB Units of Pune district. Point estimates with exact 95% confidence intervals on a logarithmic scale: bacteriologically confirmed pulmonary TB (primary endpoint; the open marker shows the leave-one-out estimate with any single AI-arm case omitted), all forms of TB, and presumptive TB pooled and by rural and urban stratum. Values above 1 favour the AI-guided arm. Source data: Table 2 and Supplementary Tables S4 and S5.

### Total case detection

Under the all-forms definition the AI arm diagnosed 15 cases and the standard campaign 32 (RR 1.26, 95% CI 0.63 to 2.39; p=0.51): Shirur 2 versus 4, Bhor 0 versus 2 and Haveli 13 versus 26. Of the standard campaign’s 32 cases, 9 were bacteriologically confirmed pulmonary TB, 5 were pulmonary TB diagnosed on chest X-ray or other clinical grounds and 18 were extrapulmonary. That campaign performed 444 chest X-rays among its 746 presumptive cases and confirmed mainly by sputum microscopy (NAAT in 95), whereas the AI sweep had no mobile X-ray, its protocol specified NAAT for every presumptive, and 4 of its 15 cases were diagnosed radiologically after referral to a facility (Supplementary Table S2).

### Presumptive yield and referral precision

The presumptive rate was 0.49% in the AI arm versus 0.42% in the standard campaign (RR 1.17, 95% CI 1.03 to 1.34; p=0.018). The difference was confined to the rural units (0.79% versus 0.36%; RR 2.22, 95% CI 1.62 to 3.00; p<0.001) and absent in the urban units (0.46% versus 0.46%; RR 1.01, 95% CI 0.87 to 1.17; p=0.94). Where the urban advantage appeared was in referral precision: 3.3% of tested presumptives in the urban AI arm (9 of 271) were bacteriologically confirmed pulmonary TB, against 1.3% in the urban standard campaign (7 of 523), a 2.5-fold difference (95% CI 0.93 to 6.59; Fisher p=0.067); pooled across the four units the proportions were 3.1% and 1.2% (10 of 325 versus 9 of 746; 2.6-fold, 95% CI 1.05 to 6.22; p=0.043). The all-forms proportions were similar (urban 4.8% versus 5.0%; pooled 4.6% versus 4.3%).

### Efficiency and cost

The AI arm screened 6,610 people per confirmed pulmonary case against 19,702 in the standard campaign (66% fewer). At the programme’s ACF unit cost of about Rs 140 per person screened, cost per confirmed case fell from Rs 27.6 lakh to Rs 9.3 lakh (66% lower).

### Acceptability and usability

The 100 FGD participants were 86 ASHAs with supervisory staff, multipurpose workers and TB health visitors; 32 had more than 10 years of field experience, all owned smartphones, 87% used Google Maps and all preferred Marathi-language outputs. In 12 of the 16 groups participants said they would use the plan in their next campaign (two further groups were split about 70:30 in favour, one was undecided and one, in a dense urban PCMC area, was opposed); 15 of 16 would recommend it to colleagues; 12 of 16 scored its usefulness at the maximum of 5, with a group median of 5 for both usefulness and ease of use; and 15 of 16 rated it better than the paper-based method. Two problems recurred. The people-to-screen target shown in each plan was endorsed by only 3 of 16 groups (19%), who read it as a quota on an already heavy workload, and turn-by-turn navigation failed for 5 of 7 workers in one dense urban PMC group. In the time- and-motion study the five plan-access steps, spread across three separate applications, took a median of 54 seconds per hotspot (IQR 42 to 69) among the 37 workers for whom every step was timed.

In the post-campaign survey (n=101; Table 3), 96% reported finding the same presumptive cases while screening fewer people (60% fully, 36% partly), 94% found navigation easy, 92% reported reduced workload, 86% time saved and 98% adequate training; 83% agreed the hotspots were higher-risk areas, but only 41% perceived more presumptive cases in hotspots than in general areas. Field barriers were also reported: 62% still encountered stigma, 37% found it very difficult to persuade people to attend for testing, and only 49% said referred persons gave sputum on time (Supplementary Table S8). Table 4 integrates the quantitative and qualitative findings.

**Table 3.** Post-campaign survey of ASHAs who used AI-NETRA microplans (n=101; 3 to 30 June 2026)

| Statement | Agreed, n (%) | Of which fully or strongly, n (%) |
|---|---|---|
| Found the same presumptive cases while screening fewer people | 97 (96) | 61 (60) |
| Adequately informed and trained for the sweep | 99 (98) | two-option item |
| Navigation to the mapped hotspots was easy | 95 (94) | 50 (50) |
| Daily workload reduced compared with previous campaigns | 93 (92) | 57 (56) |
| Field time saved | 87 (86) | 49 (49) |
| Referral to the diagnostic centre named in the plan was easy | 87 (86) | 31 (31) |
| Hotspots identified by AI-NETRA are higher-risk areas | 84 (83) | 48 (48) |
| More presumptive cases found in hotspots than in general areas | 41 (41) | three-option item |

Response options and counts for every item, including items on field barriers, are given in Supplementary Table S8. Eight respondents reported working in non-hotspot areas; excluding them changes no percentage by more than one point.

**Table 4.**
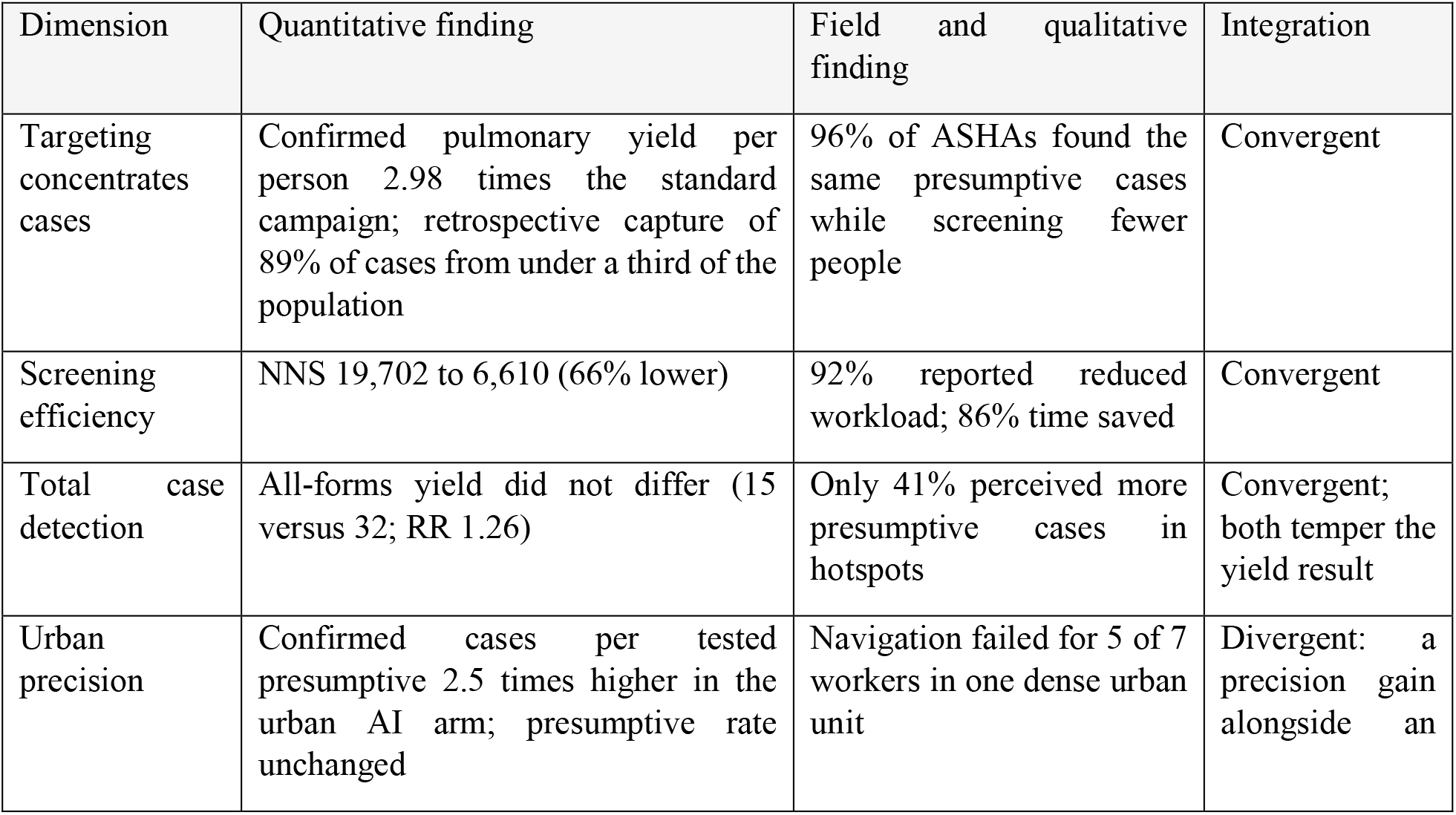
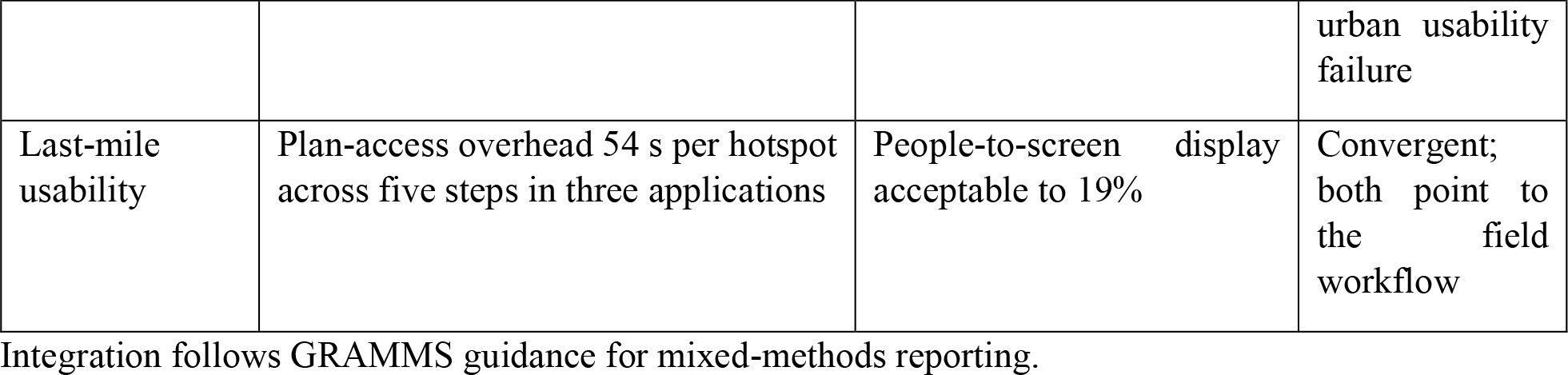
Joint display of quantitative and qualitative findings.

## Discussion

Three conclusions follow. First, the operational case is made: AI-guided planning concentrated screening into pockets with about three times the standard campaign’s confirmed yield per person, cut NNS and cost per confirmed case by two-thirds, and was accepted by the workers who used it. Second, superiority in total case detection is not established: the all-forms comparison is a null, and in absolute terms the standard campaign found twice as many cases. Third, the primary effect is real but fragile and confounded, and should be read as hypothesis-generating.

The two yield measures are reconciled by the study conditions rather than by the targeting. The standard campaign ran with a mobile X-ray van and detected clinically diagnosed and extrapulmonary disease that a symptom screen with sputum NAAT cannot: 23 of its 32 cases were X-ray-based pulmonary or extrapulmonary diagnoses, against 5 of the AI arm’s 15, a profile consistent with the large subclinical fraction of TB now recognised in prevalence surveys.^[17,18]^ Ascertainment also differed in the opposite direction. The AI arm’s protocol specified NAAT for every presumptive, whereas the standard campaign relied mainly on sputum microscopy, so part of the per-person bacteriological advantage may reflect the more sensitive test rather than better targeting. Half of the surveyed ASHAs reported that referred persons did not give sputum on time, so sputum-based confirmation without imaging was itself a constraint on what the AI arm could detect. The comparator was drawn from an earlier period, and urban denominators were constructed differently in the two arms; reconstructing the AI arm’s urban denominator from households visited lowers the pooled rate ratio to 2.71 (95% CI 0.99 to 7.54). None of these is a property of the algorithm, and none can be separated from it in this design.

The primary advantage arose in the urban units, where nine of the ten confirmed cases were found. There it took the form of precision rather than volume: the presumptive rate did not rise, but a tested presumptive in the AI arm was 2.5 times as likely to be bacteriologically confirmed. This is the setting where administrative vulnerability layers offer least, since they resolve to the block or ward and a whole-ward campaign inside a dense city is not feasible.^[10]^ Sub-ward targeting complements such layers, which allocate effort across a district, rather than replacing them. The usability failures were also urban: navigation failed in one dense unit, and the plan-access overhead fell on exactly these teams. Within the urban arm, Haveli PMC yielded 7 cases from 27,047 screened and Haveli PCMC 2 from 32,240; whether that reflects implementation fidelity or underlying epidemiology cannot be resolved here.

Two further points bear on deployment. A notification-trained model is weakest where notifications are sparse, which are often the least-served areas, so an environmental cold-start layer is needed before such targeting can claim to reach them. And prioritisation alone does not raise total detection: the all-forms null is the expected result of prioritising without then saturating the prioritised zones with complete coverage, imaging and molecular testing, the approach that community-wide screening has been shown to require to reduce prevalence.^[19]^ AI-NETRA is best read as the first layer of that sequence. Earlier geospatial TB work has largely operated at district or province level without field validation,^[7,20]^ and community ACF reviews emphasise that yield depends as much on the diagnostic package as on where teams go.^[21]^

Strengths include a pre-specified bacteriological endpoint applied to both arms, programme conditions, a mixed-methods design and full reporting of the null. The limitations are substantial: a non-concurrent comparator exposed to secular and seasonal trends; unmatched case ascertainment in both directions; population-covered urban denominators; an intervention bundled with training and a new workflow; small counts, so that the primary result loses conventional significance with the omission of a single case; a single district; self-reported acceptability subject to social desirability, with FGDs facilitated by implementation-partner staff and documented by note-taking rather than transcription; and manual geolocation of cases, which limits within-hexagon spatial analysis.

The decisive next study is a concurrent cluster-randomised or stepped-wedge trial in which both arms run simultaneously with harmonised case definitions, equal access to X-ray and NAAT and equivalent training, with co-primary outcomes of total cases found and cases missed in deprioritised areas, sited in a low-notification district and including a dense urban arm. Routine export of the geo-coordinates already held in Nikshay would allow hexagon-level attribution without bespoke geocoding.

## Conclusion

AI-guided planning raised bacteriologically confirmed yield per person screened about threefold, lowered NNS and cost per confirmed case by two-thirds and was well received by frontline workers, but total case detection did not differ from a standard campaign that had mobile X-ray and ran at a different time. The efficiency case for geospatial targeting as a layer within ACF is strong, and strongest in dense urban settings; the case that it finds more cases in absolute terms requires a concurrent, like-for-like trial.

## Supporting information

Supplementary Code (statistical analysis script)

Supplementary Data (workbook, Tables S1 to S10)

Supplementary Appendix (methods, results, TREND and COREQ checklists)

## Data Availability

Aggregate data underlying every table and figure are provided in the Supplementary Data workbook. An open-source implementation of the AI-NETRA ACF model (feature construction, the two-stage hurdle model, evaluation and ranked-hexagon output, with a synthetic-data generator) is available under the Apache-2.0 licence at https://github.com/Froncort/AI-ACF-ML-Algorithm. The repository contains the model and evaluation code with a synthetic-data generator; it does not include patient data, fitted model weights, or the platform used to ingest registers, geocode addresses and generate the field plans. The statistical analysis code for this paper is provided as Supplementary Data alongside the workbook. Individual-level notification data used for model training and individual-level programme data are held by the Maharashtra State TB Office under NTEP data governance and are not publicly available. The retrospective validation was presented as an abstract at the Union World Conference on Lung Health 2026; the complete pilot study report (22 July 2026) was released to the State TB Office and is published at https://www.froncort.ai/ai-netra/pilot-study (accessed 9 November 2026).

https://www.froncort.ai/ai-netra/pilot-study

## Declarations

### Contributors

UR conceived the study, led the development and deployment of the tool, performed the statistical analysis and drafted the manuscript. HV led field implementation and data acquisition and contributed to the FGD analysis. MD analysed, summarised and reported the FGDs. RP provided programme direction, data access and interpretation. SC provided institutional leadership of the study at FIND and oversaw the implementation partner’s field operations. AB was principal investigator at FIND, conceived and supervised the study and interpreted the findings. All authors revised the manuscript critically, approved the final version and agree to be accountable for the work.

## Acknowledgements

We thank Mahesh Kolle (SHAiPE Foundation, Pune) for field coordination and data collection; Himanshu Gholse, Aayush Agrawal, Pranav Metil, Parth Manekar, Sara Nambiar, Isha Shah (Froncort.AI) for platform development and plan generation; the District TB Officers and staff of Pune Rural, Pune Municipal Corporation and Pimpri-Chinchwad Municipal Corporation; the ASHAs, Supervisors, TB health visitors and field staff who conducted the screening; and the participants of the focus groups and survey.

## Ethics

The Ethics Committee of the Jehangir Clinical Development Centre, Pune, India, gave ethical approval for this work (reference JCDC/BHR/26/005, 13 March 2026). The prospective field evaluation was conducted under this approval. Routine programme data of the National Tuberculosis Elimination Programme were accessed by FIND under a non-disclosure agreement with the State TB Office, Government of Maharashtra, and shared with Froncort.AI under a back-to-back non-disclosure agreement between FIND and Froncort.AI; these agreements govern the use of the data in accordance with the programme’s data-governance arrangements and India’s Digital Personal Data Protection Act, 2023.

## Registration

The study evaluated a planning method within routine programme operations: individuals received the standard NTEP screening and diagnostic pathway, the intervention determined only which areas field teams visited, and the comparator was a completed programme campaign. It was therefore not registered as a clinical trial.

## Data and code availability

Aggregate data underlying every table and figure are provided in the Supplementary Data workbook. An open-source implementation of the AI-NETRA ACF model (feature construction, the two-stage hurdle model, evaluation and ranked-hexagon output, with a synthetic-data generator) is available under the Apache-2.0 licence at https://github.com/Froncort/AI-ACF-ML-Algorithm.^[22]^ The repository contains the model and evaluation code with a synthetic-data generator; it does not include patient data, fitted model weights, or the platform used to ingest registers, geocode addresses and generate the field plans. The statistical analysis code for this paper is provided as Supplementary Data alongside the workbook. Individual-level notification data used for model training and individual-level programme data are held by the Maharashtra State TB Office under NTEP data governance and are not publicly available.

## Prior dissemination

The retrospective validation was presented as an abstract at the Union World Conference on Lung Health 2026; the complete pilot study report (22 July 2026) was released to the State TB Office and is published at https://www.froncort.ai/ai-netra/pilot-study (accessed 9 November 2026).

## Use of generative AI

This manuscript was derived from the authors’ study report and statistical analysis plan. The authors used Claude (Anthropic), a large language model-based assistant, to restructure that material into journal format, to edit the text for length and clarity, to recompute the reported statistics from the source data as a verification step, and to format the tables, the Supplementary Data workbook and Figure 1. The study design, data collection, analyses, interpretation and conclusions are the authors’ own; the assistant was not used to generate, impute or alter any study data, and no image content was generated by AI. The authors reviewed and revised all text, checked every figure against the source records and take full responsibility for the content. AI-NETRA, the system evaluated in this study, is a machine-learning planning tool described in Methods and is distinct from the writing assistant. Generative AI tools do not meet authorship criteria and none is listed as an author.

