## Supplementary Code (statistical analysis script) for "Geospatial AI targeting of tuberculosis active case finding in Pune, India: a quasi-experimental evaluation"

Geospatial AI targeting of tuberculosis active case finding in Pune, India: a quasi-experimental evaluation. File: analysis_code_AI_NETRA_prospective.py (Python 3.10+, SciPy 1.11, NumPy). The script reproduces every statistic in the manuscript, the Supplementary Appendix and the Supplementary Data workbook from the aggregate counts it contains; the same file is supplied in plain text.

```
#!/usr/bin/env python3
"""
Statistical analysis code for:
  Geospatial AI targeting of tuberculosis active case finding in Pune, India:
  a quasi-experimental evaluation (AI-NETRA prospective field evaluation).

Reproduces every statistic reported in the manuscript, the Supplementary
Appendix and the Supplementary Data workbook from the aggregate counts below.

Inputs (aggregate counts, as in Supplementary Tables S1 to S3):
  AI-guided arm, field data extract 29 June 2026, by TB Unit:
      households visited, persons screened, presumptive TB, tested,
      all-forms cases, NAAT-positive pulmonary cases (primary endpoint).
  Standard arm, district TB-4 compilation, ACF campaign Dec 2024 to Jan 2025:
      persons screened, presumptive TB, sputum smear- or NAAT-positive
      pulmonary TB (primary comparator), clinically diagnosed pulmonary TB,
      extrapulmonary TB.

Methods:
  Yield per 10,000 = cases / persons screened x 10,000.
  Exact two-sided 95% Poisson (Garwood) interval for a count k:
      lower = chi2.ppf(0.025, 2k) / 2 (0 when k = 0), upper = chi2.ppf(0.975, 2k + 2) / 2.
  NNS = persons screened / cases; interval = persons screened / (upper, lower).
  Rate ratio between arms: exact conditional (binomial) test. Conditional on
      a + b = n, a ~ Binomial(n, p) with p = RR t1 / (RR t1 + t2); two-sided
      exact p-value tests p0 = t1 / (t1 + t2); Clopper-Pearson limits for p are
      transformed to RR = p / (1 - p) x t2 / t1.
  Presumptive yield and referral precision: Fisher's exact test on the 2 x 2
      table, Katz log-normal interval for the ratio of proportions.
  Leave-one-out: each AI-arm case omitted in turn (all omissions identical).
  Denominator sensitivity: urban AI-arm persons screened reconstructed as
      households visited x persons per household observed in the rural units.

Requirements: Python 3.10+, scipy >= 1.11, numpy.
Run:  python analysis_code_AI_NETRA_prospective.py
"""

from __future__ import annotations

import csv
import sys
from dataclasses import dataclass

import numpy as np
from scipy.stats import binomtest, chi2, fisher_exact

# ---------------------------------------------------------------------------
# Inputs
# ---------------------------------------------------------------------------

AI_ARM = {
    # unit: (households, screened, presumptive, tested, all_forms, primary)
    "Shirur": (855, 3710, 31, 31, 2, 1),
    "Bhor": (815, 3098, 23, 23, 0, 0),
    "Haveli PMC": (7004, 27047, 204, 204, 9, 7),
    "Haveli PCMC": (9143, 32240, 68, 67, 4, 2),
}

STANDARD_ARM = {
```

```
    # unit: (screened, presumptive, bact_pulm, clinical_pulm, eptb, naat_done, xray_done)
    "Bhor": (19842, 81, 1, 0, 1, 5, 11),
    "Shirur": (42664, 142, 1, 1, 2, 30, 114),
    "Haveli AFMC": (64730, 207, 5, 3, 5, 30, 180),
    "Haveli BJMC": (50086, 316, 2, 1, 10, 30, 139),
}

UNIT_COST_RS_PER_PERSON = 140.0  # programme ACF unit cost per person screened


# ---------------------------------------------------------------------------
# Methods
# ---------------------------------------------------------------------------

def garwood(k: int) -> tuple[float, float]:
    """Exact two-sided 95% Poisson interval for an observed count k."""
    lo = 0.0 if k == 0 else chi2.ppf(0.025, 2 * k) / 2
    hi = chi2.ppf(0.975, 2 * k + 2) / 2
    return lo, hi


@dataclass
class RateRatio:
    rr: float
    lo: float
    hi: float
    p: float

    def fmt(self, nd: int = 2) -> str:
        hi = "inf" if np.isinf(self.hi) else f"{self.hi:.{nd}f}"
        return f"{self.rr:.{nd}f} (95% CI {self.lo:.{nd}f} to {hi}); p={self.p:.3f}"


def exact_rate_ratio(a: int, t1: float, b: int, t2: float) -> RateRatio:
    """Exact conditional binomial rate ratio of arm 1 (a cases in t1) to arm 2 (b in t2)."""
    n = a + b
    p0 = t1 / (t1 + t2)
    rr = (a / t1) / (b / t2) if b > 0 else float("inf")
    test = binomtest(a, n, p0, alternative="two-sided")
    ci = test.proportion_ci(confidence_level=0.95, method="exact")
    lo = ci.low / (1 - ci.low) * t2 / t1 if ci.low > 0 else 0.0
    hi = ci.high / (1 - ci.high) * t2 / t1 if ci.high < 1 else float("inf")
    return RateRatio(rr, lo, hi, test.pvalue)


def katz_ratio(a: int, n1: int, b: int, n2: int) -> tuple[float, float, float, float]:
    """Ratio of proportions a/n1 : b/n2 with Katz 95% interval and Fisher exact p."""
    rr = (a / n1) / (b / n2)
    se = np.sqrt(1 / a - 1 / n1 + 1 / b - 1 / n2)
    lo, hi = np.exp(np.log(rr) - 1.96 * se), np.exp(np.log(rr) + 1.96 * se)
    p = fisher_exact([[a, n1 - a], [b, n2 - b]])[1]
    return rr, lo, hi, p


def per10k(k: int, t: float) -> float:
    return k / t * 1e4


# ---------------------------------------------------------------------------
# Analysis
# ---------------------------------------------------------------------------

def main(out_csv: str | None = "analysis_output.csv") -> None:
    rows: list[dict] = []

    def record(section: str, item: str, value: str) -> None:
        rows.append({"section": section, "item": item, "value": value})
        print(f"{section:22s} | {item:58s} | {value}")
```

```
    # Totals
    ai_hh = sum(v[0] for v in AI_ARM.values())
    ai_scr = sum(v[1] for v in AI_ARM.values())
    ai_pres = sum(v[2] for v in AI_ARM.values())
    ai_tested = sum(v[3] for v in AI_ARM.values())
    ai_all = sum(v[4] for v in AI_ARM.values())
    ai_prim = sum(v[5] for v in AI_ARM.values())

    st_scr = sum(v[0] for v in STANDARD_ARM.values())
    st_pres = sum(v[1] for v in STANDARD_ARM.values())
    st_bact = sum(v[2] for v in STANDARD_ARM.values())
    st_clin = sum(v[3] for v in STANDARD_ARM.values())
    st_eptb = sum(v[4] for v in STANDARD_ARM.values())
    st_all = st_bact + st_clin + st_eptb
    st_naat = sum(v[5] for v in STANDARD_ARM.values())
    st_xray = sum(v[6] for v in STANDARD_ARM.values())

    record("Cascade", "AI arm households / screened / presumptive / tested",
           f"{ai_hh:,} / {ai_scr:,} / {ai_pres} ({ai_pres/ai_scr*100:.2f}%; {ai_pres/ai_hh*100:.2f} per 100 hh)
/ {ai_tested} ({ai_tested/ai_pres*100:.1f}%)")
    record("Cascade", "AI arm all-forms / primary cases", f"{ai_all} ({ai_all/ai_tested*100:.1f}% of tested) /
{ai_prim}")
    record("Cascade", "Standard arm screened / presumptive / bact pulm / clinical pulm / EPTB / total",
           f"{st_scr:,} / {st_pres} / {st_bact} / {st_clin} / {st_eptb} / {st_all}; NAAT {st_naat}; X-ray {st_x
ray}")
    record("Cascade", "AI screened as % of standard", f"{ai_scr/st_scr*100:.1f}%")

    # Table 1: unit yields and NNS
    for unit, (hh, scr, pres, tested, allf, prim) in AI_ARM.items():
        lo, hi = garwood(prim)
        nns = f"{scr/prim:,.0f}" if prim else f">{scr/hi:,.0f} (lower bound)"
        record("Table 1", f"{unit}: primary yield per 10,000 (95% CI); NNS",
               f"{per10k(prim, scr):.2f} ({per10k(lo, scr):.2f} to {per10k(hi, scr):.2f}); {nns}")
    lo, hi = garwood(ai_prim)
    record("Table 1", "Pooled primary yield per 10,000 (95% CI)",
           f"{per10k(ai_prim, ai_scr):.2f} ({per10k(lo, ai_scr):.2f} to {per10k(hi, ai_scr):.2f}); Garwood even
t limits {lo:.2f} to {hi:.2f}")
    record("Table 1", "Pooled NNS (95% CI)", f"{ai_scr/ai_prim:,.0f} ({ai_scr/hi:,.0f} to {ai_scr/lo:,.0f})")
    record("Table 2", "Standard arm primary yield per 10,000; NNS", f"{per10k(st_bact, st_scr):.3f}; {st_scr/st
_bact:,.0f}")
    record("Table 2", "NNS reduction", f"{(1 - (ai_scr/ai_prim)/(st_scr/st_bact))*100:.1f}%")

    # Table 2: rate ratios
    st_unit = {"Bhor": STANDARD_ARM["Bhor"], "Shirur": STANDARD_ARM["Shirur"]}
    hav_scr = STANDARD_ARM["Haveli AFMC"][0] + STANDARD_ARM["Haveli BJMC"][0]
    hav_bact = STANDARD_ARM["Haveli AFMC"][2] + STANDARD_ARM["Haveli BJMC"][2]
    hav_all = sum(STANDARD_ARM[u][2] + STANDARD_ARM[u][3] + STANDARD_ARM[u][4] for u in ("Haveli AFMC", "Haveli
BJMC"))
    ai_hav_scr = AI_ARM["Haveli PMC"][1] + AI_ARM["Haveli PCMC"][1]
    ai_hav_prim = AI_ARM["Haveli PMC"][5] + AI_ARM["Haveli PCMC"][5]
    ai_hav_all = AI_ARM["Haveli PMC"][4] + AI_ARM["Haveli PCMC"][4]

    comparisons = {
        "Primary, Bhor": (AI_ARM["Bhor"][5], AI_ARM["Bhor"][1], st_unit["Bhor"][2], st_unit["Bhor"][0]),
        "Primary, Shirur": (AI_ARM["Shirur"][5], AI_ARM["Shirur"][1], st_unit["Shirur"][2], st_unit["Shirur"]
[0]),
        "Primary, Haveli (PMC and PCMC)": (ai_hav_prim, ai_hav_scr, hav_bact, hav_scr),
        "Primary, pooled": (ai_prim, ai_scr, st_bact, st_scr),
        "Primary, pooled, any one AI-arm case omitted": (ai_prim - 1, ai_scr, st_bact, st_scr),
        "Primary, pooled, any one standard-arm case omitted": (ai_prim, ai_scr, st_bact - 1, st_scr),
        "Primary, rural (Bhor and Shirur)": (1, AI_ARM["Bhor"][1] + AI_ARM["Shirur"][1], 2, st_unit["Bhor"][0]
+ st_unit["Shirur"][0]),
        "All forms, Bhor": (AI_ARM["Bhor"][4], AI_ARM["Bhor"][1], sum(st_unit["Bhor"][2:5]), st_unit["Bhor"]
[0]),
        "All forms, Shirur": (AI_ARM["Shirur"][4], AI_ARM["Shirur"][1], sum(st_unit["Shirur"][2:5]), st_unit["S
hirur"][0]),
        "All forms, Haveli (PMC and PCMC)": (ai_hav_all, ai_hav_scr, hav_all, hav_scr),
```

```
        "All forms, pooled": (ai_all, ai_scr, st_all, st_scr),
        "Sensitivity: Nikshay flag at face value (11 vs 9)": (11, ai_scr, st_bact, st_scr),
    }
    for name, (a, t1, b, t2) in comparisons.items():
        record("Table 2 / S4 / S6", f"{name}: {a}/{t1:,} vs {b}/{t2:,}", exact_rate_ratio(a, t1, b, t2).fmt())

    # Table S5: presumptive yield and referral precision
    rural_ai = (AI_ARM["Bhor"][2] + AI_ARM["Shirur"][2], AI_ARM["Bhor"][1] + AI_ARM["Shirur"][1])
    rural_st = (st_unit["Bhor"][1] + st_unit["Shirur"][1], st_unit["Bhor"][0] + st_unit["Shirur"][0])
    urban_ai = (AI_ARM["Haveli PMC"][2] + AI_ARM["Haveli PCMC"][2], ai_hav_scr)
    urban_st = (STANDARD_ARM["Haveli AFMC"][1] + STANDARD_ARM["Haveli BJMC"][1], hav_scr)
    for name, (a, n1), (b, n2) in [("Pooled", (ai_pres, ai_scr), (st_pres, st_scr)),
                                   ("Rural", rural_ai, rural_st), ("Urban", urban_ai, urban_st)]:
        rr, lo, hi, p = katz_ratio(a, n1, b, n2)
        record("Table S5 presumptive", f"{name}: {a}/{n1:,} ({a/n1*100:.2f}%) vs {b}/{n2:,} ({b/n2*100:.2f}%)",
               f"ratio {rr:.2f} (Katz 95% CI {lo:.2f} to {hi:.2f}); Fisher p={p:.4f}")

    urban_tested_ai = AI_ARM["Haveli PMC"][3] + AI_ARM["Haveli PCMC"][3]
    precision = {
        "Urban, primary definition": (ai_hav_prim, urban_tested_ai, hav_bact, urban_st[0]),
        "Pooled, primary definition": (ai_prim, ai_tested, st_bact, st_pres),
        "Urban, all forms": (ai_hav_all, urban_tested_ai, hav_all, urban_st[0]),
        "Pooled, all forms": (ai_all, ai_tested, st_all, st_pres),
    }
    for name, (a, n1, b, n2) in precision.items():
        rr, lo, hi, p = katz_ratio(a, n1, b, n2)
        record("Table S5 precision", f"{name}: {a}/{n1} ({a/n1*100:.2f}%) vs {b}/{n2} ({b/n2*100:.2f}%)",
               f"ratio {rr:.2f} (Katz 95% CI {lo:.2f} to {hi:.2f}); Fisher p={p:.3f}")

    # Cost
    for label, nns in [("AI arm", ai_scr / ai_prim), ("Standard arm", st_scr / st_bact)]:
        record("Table S7 cost", f"{label}: cost per confirmed case at Rs {UNIT_COST_RS_PER_PERSON:.0f} per pers
on",
               f"Rs {nns*UNIT_COST_RS_PER_PERSON/1e5:.1f} lakh (NNS {nns:,.0f})")

    # Denominator sensitivity (Table S6)
    rural_hh = AI_ARM["Bhor"][0] + AI_ARM["Shirur"][0]
    rural_scr = AI_ARM["Bhor"][1] + AI_ARM["Shirur"][1]
    urban_hh = AI_ARM["Haveli PMC"][0] + AI_ARM["Haveli PCMC"][0]
    ratio_rural = rural_scr / rural_hh
    ratio_overall = ai_scr / ai_hh
    record("Table S6 denominators", "Persons per household: rural / urban as recorded / overall",
           f"{ratio_rural:.3f} / {ai_hav_scr/urban_hh:.3f} / {ratio_overall:.3f}")
    for label, ratio in [("rural ratio", ratio_rural), ("overall ratio", ratio_overall)]:
        urban_rec = urban_hh * ratio
        pooled_rec = rural_scr + urban_rec
        record("Table S6 denominators", f"Urban AI denominator reconstructed at the {label} ({ratio:.2f})",
               f"urban {urban_rec:,.0f}, pooled {pooled_rec:,.0f}; urban RR {exact_rate_ratio(ai_hav_prim, roun
d(urban_rec), hav_bact, hav_scr).fmt()}; "
               f"pooled RR {exact_rate_ratio(ai_prim, round(pooled_rec), st_bact, st_scr).fmt()}; pooled NNS {p
ooled_rec/ai_prim:,.0f}")

    if out_csv:
        with open(out_csv, "w", newline="", encoding="utf-8") as fh:
            w = csv.DictWriter(fh, fieldnames=["section", "item", "value"])
            w.writeheader()
            w.writerows(rows)
        print(f"\nWritten: {out_csv}")


if __name__ == "__main__":
    main(sys.argv[1] if len(sys.argv) > 1 else "analysis_output.csv")
```
