## Supplementary Appendix (methods, results, TREND and COREQ checklists) for "Geospatial AI targeting of tuberculosis active case finding in Pune, India: a quasi-experimental evaluation"

Supplementary Appendix to: Geospatial AI targeting of tuberculosis active case finding in Pune, India: a quasi-experimental evaluation. Rao U, Pawar R, Beri A, Chadha S, Vashistha H, Das M. Supplementary Data (workbook and analysis code) are provided as separate files.

### S1. Supplementary methods

#### S1.1 The planning tool

AI-NETRA (version 1) is a geospatial, agentic decision-support system that scores TB risk per H3 hexagon and assembles the highest-scoring hexagons into field microplans. Training data were the Pune Rural Nikshay notification register for 2022 to 2025, restricted to bacteriologically confirmed cases (actively and passively detected; about 98% passively detected) and geocoded to residential address with an external geocoding service. Geocoding assigned about 92% of records to hexagons; 16% resolved to building precision, 12% to geometric centre, 1% to an interpolated range and 72% to area precision. All geocoded records were retained because restricting to building-precision addresses would have left too few for spatial learning. Hexagons are scored on five signal domains: notified cases in the preceding 1, 3 and 6 months with temporal decay; case characteristics, with pulmonary disease weighted higher for transmissibility; spatial spillover from adjacent hexagons; cold-start inference from population density and socioeconomic proxies where notification history is sparse; and programmatic vulnerability indicators (age and comorbidity profiles of notified cases). Scores in the top 10% define high-priority hexagons and the 10th to 30th percentile medium-priority hexagons. The scoring model is a two-stage gradient-boosted (XGBoost) hurdle: a gate classifier estimates the probability that a hexagon notifies any case in the following month, isotonic regression calibrates that probability, and a Poisson count model estimates the expected number of cases when the gate is open; the product of the two ranks hexagons, with features computed at month M predicting notifications at month M+1. An open-source implementation with a synthetic-data walkthrough is available under the Apache-2.0 licence (main text, Data and code availability); it assigns tiers by a count-and-probability rule rather than the percentile rule used in the deployed plans, and its optional contact-tracing features were not used in the Pune model.

Navigation hotspots are provided at H3 resolution 10 (hexagon area about 0.015 km², equivalent to a 123 x 123 m square); the screening population and NNS denominator are computed at resolution 9 (about 0.105 km², a 325 x 325 m square), because a resolution-10 cell can imply screening a handful of people in a neighbourhood of hundreds, which field teams cannot action. Each plan lists, for every hotspot: a landmark name; a street address and map link; the residential clusters within the cell; cases notified in the preceding six months; a predicted case range; and the number of people to screen. Plans were delivered as PDF documents with map links. A single locked model version generated all plans before the sweep; no retraining occurred during the field phase.

#### S1.2 Microplan inventory and sampling

Table S1 (Supplementary Data workbook) lists, for each unit, the number of high- and medium-priority hexagons and the planned screening population in the district plan from which the four units were selected, beside the population actually screened. Coverage rules were: every high-priority hexagon screened completely; about one-third of medium-priority hexagons, chosen by rank order within geographically stratified groups so that no part of a unit was left unsampled; and non-hotspot (low-priority) areas adjacent to planned hexagons screened opportunistically as a specificity check. The rural units screened more people than their planned hotspot population because teams screened whole settlement clusters around each hotspot; the urban units screened about half of the planned population under the one-third rule for medium-priority cells.

#### S1.3 Statistical methods

Yield per 10,000 screened is cases divided by persons screened. Exact two-sided 95% intervals for a Poisson count k use the Garwood (chi-square) method: lower limit = $\chi^2(0.025, 2k)/2$ and upper limit = $\chi^2(0.975, 2k + 2)/2$, with the lower limit 0 when k = 0. NNS = persons screened divided by cases, with the interval obtained by dividing persons screened by the upper and lower Poisson limits. For two arms with a and b cases over t1 and t2 persons screened, the rate ratio RR = (a/t1)/(b/t2); conditional on a + b = n, a follows a binomial distribution with success probability p = RR·t1/(RR·t1 + t2); the exact two-sided p-value tests p0 = t1/(t1 + t2), and the Clopper-Pearson interval for p is transformed to RR = p/(1 − p) · t2/t1. Presumptive yield used Fisher's exact test on the 2 x 2 table of presumptive versus not presumptive by arm, with log-binomial regression for the rate ratio and its interval. Leave-one-out sensitivity recomputed the primary rate ratio after omitting each confirmed case in turn. All exact computations were repeated independently for this manuscript with SciPy 1.11 (scipy.stats.binomtest, two-sided; beta and chi-square quantiles) and agree with the locked study report to the precision shown.

#### S1.4 Costing basis

Cost per confirmed case = NNS multiplied by the programme's ACF unit cost per person screened. The unit cost implied by the locked figures (Rs 27.6 lakh per case at NNS 19,702; Rs 9.3 lakh at NNS 6,610) is about Rs 140 per person screened, the programme's ACF cost per person screened as applied in the study report; its components are not itemised here. Because the same unit cost is applied to both arms, the percentage reduction in cost per case equals the percentage reduction in NNS; no cost of the planning tool itself is included.

#### S1.5 Acceptability and usability instruments

Focus group discussions: 16 groups across the four units (Shirur 3, Bhor 5, Haveli PMC 4, Haveli PCMC 4) held on 17 April (Shirur), 22 April (Bhor), 29 April (Haveli PMC) and 11 May 2026 (Haveli PCMC), with 100 participants (86 ASHAs, 5 senior treatment supervisors, 1 senior TB laboratory supervisor, 1 multipurpose worker and 7 TB health visitors), facilitated in Marathi by the implementation partner's field staff with a moderator and a note-taker per session. Each session combined a hands-on demonstration, in which every participant opened a microplan and worked through each feature, with a structured five-part template: participant profile and digital skills; current ACF practice and the AI-NETRA plan; eight mobile-usability statements rated from 1 (strongly disagree) to 5 (strongly agree); field context, workflow integration, supervision, trust and suggestions; and a 17-item feature-priority vote (endorse or reject) with four overall acceptability questions. Ratings and votes were averaged within each group and analysed with the group as the unit of analysis; note-taker observations and investigator field notes were synthesised by content. The compiled template (18 May 2026) and the FGD analysis report (version 3.0, 4 June 2026) are the source documents. Time-and-motion: 57 workers

(Shirur 17, Bhor 8, Haveli PMC 20, Haveli PCMC 12) were timed in seconds through five steps: opening the plan PDF from WhatsApp, locating the assigned hotspot row, tapping the satellite-map link, orienting the map to the current location, and starting turn-by-turn navigation; travel and on-site screening times were not recorded, so the planned comparison with the paper workflow was not possible. Post-campaign survey: 101 ASHAs who had used microplans completed a 28-item structured questionnaire in Marathi, delivered as an online form between 3 and 30 June 2026; 93 reported having worked in AI-NETRA hotspot areas and 8 in regular areas (items in Table 3 and Supplementary Table S8).

### S2. Supplementary results

#### S2.1 Standard-arm cascade (Table S2)

The December 2024 to January 2025 campaign in the four units, from the district TB-4 compilation: 177,322 persons screened; 746 presumptive TB identified, all with samples collected; 740 sputum microscopy; 444 chest X-rays; 95 NAAT; 63 abnormal X-rays; 9 sputum- or NAAT-positive pulmonary TB; 5 pulmonary TB diagnosed by X-ray or other method; 18 extrapulmonary TB; 32 diagnosed in total and 33 recorded as put on treatment. By unit: Bhor 19,842 screened, 81 presumptive, 11 X-rays, 5 NAAT, 1 bacteriologically confirmed pulmonary, 0 clinical pulmonary, 1 extrapulmonary; Shirur 42,664 screened, 142 presumptive, 114 X-rays, 30 NAAT, 1, 1 and 2; Haveli (AFMC and BJMC units combined) 114,816 screened, 523 presumptive, 319 X-rays, 60 NAAT, 7, 4 and 15. The TB-4 form records bacteriologically confirmed pulmonary TB as sputum-smear or NAAT positive; the form records six NAAT-positive (MTB detected) diagnoses across all forms in these units, so most but not necessarily all of the nine were NAAT-confirmed and the remainder smear-positive.

#### S2.2 Presumptive yield and referral precision (Table S5)

| Stratum | AI arm presumptive / screened (%) | Standard presumptive / screened (%) | Rate ratio (95% CI) | Fisher p |
|---|---|---|---|---|
| Pooled | 326 / 66,095 (0.49) | 746 / 177,322 (0.42) | 1.17 (1.03 to 1.34) | 0.018 |
| Rural (Bhor, Shirur) | 54 / 6,808 (0.79) | 223 / 62,506 (0.36) | 2.22 (1.62 to 3.00) | <0.001 |
| Urban (Haveli) | 272 / 59,287 (0.46) | 523 / 114,816 (0.46) | 1.01 (0.87 to 1.17) | 0.94 |

Intervals from log-binomial regression (locked study report); Katz intervals computed for this manuscript are 1.03 to 1.33, 1.65 to 2.99 and 0.87 to 1.17. Referral precision, bacteriologically confirmed pulmonary TB per tested presumptive: urban AI arm 9 of 271 (3.3%) versus urban standard 7 of 523 (1.3%), ratio 2.48 (95% CI 0.93 to 6.59; Fisher p=0.067); pooled 10 of 325 (3.1%) versus 9 of 746 (1.2%), ratio 2.55 (95% CI 1.05 to 6.22; p=0.043); by unit, Haveli PMC 7 of 204 (3.4%) and Haveli PCMC 2 of 67 (3.0%). All-forms cases per tested presumptive: urban 13 of 271 (4.8%) versus 26 of 523 (5.0%); pooled 15 of 325 (4.6%) versus 32 of 746 (4.3%). Precision values are computed under the primary definition from the final field extract of 29 June 2026.

#### S2.3 Leave-one-out sensitivity (Table S4)

Each of the ten omissions of an AI-arm case gives the same result, 9 of 66,095 versus 9 of 177,322: rate ratio 2.68 (95% CI 0.94 to 7.63; p=0.058). Omitting any single standard-arm case gives 10 of 66,095 versus 8 of 177,322: rate ratio 3.35 (95% CI 1.19 to 9.78; p=0.013). The primary finding is therefore significant as observed but not robust to the loss of a single AI-arm event.

#### S2.4 Denominator sensitivity (urban units)

Urban denominators in both arms are population-covered counts. In the AI arm, persons screened per household visited was 4.08 in the rural units (6,808 persons, 1,670 households, individually enumerated) and 3.67 as recorded in the urban units (59,287 persons, 16,147 households). Reconstructing the urban AI-arm denominator as households visited multiplied by the rural ratio gives 65,826 urban and 72,634 pooled persons screened: the urban rate ratio becomes 2.24 (95% CI 0.74 to 7.09; p=0.12), the pooled rate ratio 2.71 (95% CI 0.99 to 7.54; p=0.039) and the pooled NNS 7,263 (Table S6). Reconstruction at the overall AI-arm ratio (3.71) reproduces the recorded denominators within 1%. The standard arm's urban denominators are also population-covered, and the TB-4 report carries no household counts, so only the AI arm is adjusted, which is conservative for the AI arm.

#### S2.5 Focus group participants and time-and-motion (Table S8)

Sixteen focus groups (Shirur 3, Bhor 5, Haveli PMC 4, Haveli PCMC 4), 100 participants: 86 ASHAs, 5 senior treatment supervisors, 1 senior TB laboratory supervisor, 1 multipurpose worker and 7 TB health visitors; 32 with more than 10 years of field experience and 3 to 4 who had joined within the preceding months; 100% smartphone owners using WhatsApp; 87% able to use Google Maps; 96% able to open a PDF; 100% preferring Marathi, 12% also comfortable in Hindi. Group-level acceptability (Table S9): 12 of 16 groups would use the AI-generated plan in their next campaign, 2 were split about 70:30 in favour, 1 was undecided and 1 opposed; 15 of 16 would recommend it to a colleague (1 undecided); usefulness was scored 5 of 5 by 12 groups, 4.4 by the three Shirur groups (7 participants scoring 5 and 10 scoring 4) and 2 by one PCMC group, a group median of 5; ease of use was scored 5 by 12 groups, 4.5 by three and 4 by one (median 5); 14 groups rated the tool much better than the paper-based method, 1 better and 1 worse. The FGD analysis report (version 3.0) summarises the same data as about 75% of groups willing to use, about 96% willing to recommend, 81% scoring usefulness at 5, and mean scores of 4.5 for usefulness and ease of use; the group counts above are taken directly from the compiled template. On the eight mobile-usability statements, 10 to 16 of the 16 groups gave the maximum score of 5; the exception was turn-by-turn navigation, which failed for 5 of the 7 workers in one Haveli PMC group (5 strongly disagree, 1 neutral, 1 agree). In the feature-priority vote, every group endorsed a priority label with colour, residential region names inside each hotspot, the tap-to-open satellite view, a full plan in Marathi, a larger font option and WhatsApp delivery; 15 endorsed offline map download and a dedicated app; 13 voice directions in Marathi, automatic advance to the next hotspot and audio read-aloud; 12 a predicted case range per hotspot and marking a hotspot done on the phone; 11 a photograph at each hotspot; 10 the historical case count and the estimated walking time; and 3 the people-to-screen number. Themes from the note-taker observations: ACF plans are issued top-down from the medical officer and senior treatment supervisor, with no worker role in area selection; workloads of 20 to 40 households a day over a fixed 14-day window; parallel registers across programmes ("80% of working time is spent on writing and maintaining multiple registers; only 20% is actual fieldwork", an ASHA); refusals to give sputum, more often by men, with occasional requests for payment; incentives unpaid since December 2025 and a mobile-data allowance of Rs 300 a month against a need of 1.5 to 2.5 GB a day; the people-to-screen and predicted-case numbers read as under-estimates of the population and as a source of anxiety if not achieved; and requests to add slums, migrant and industrial pockets, landmarks and real-time addition of locations, an offline Marathi app, counselling videos for stigma, and integration with Nikshay. Time-and-motion (n=57): the five plan-access steps took a median of 54 seconds per hotspot (IQR 42 to 69) among the 37 workers with every step timed, and 49 seconds (IQR 40 to 69) across all 57 when summed over the steps recorded; per-step medians were 8 seconds to open the PDF, 9 to locate the hotspot row, 7 to open the satellite link, 9 to orient the map and 12 to start navigation; the 20 incomplete records most often lacked the navigation step. The study report gave an interquartile range of 43 to 85 seconds for the same median; the values here are recomputed from the compiled dataset.

#### S2.6 Statistical power (Table S9)

Approximate total confirmed cases required for 80% power at two-sided alpha 0.05, from the statistical analysis plan: rate ratio 1.25, about 640; 1.5, about 200; 2.0, about 75; 3.0, about 35. The exact 95% Poisson interval around an observed count of 10 is 4.80 to 18.39.

#### S2.7 Characteristics of the AI-arm cases (Table S3b)

| Characteristic | All forms (n=15) | Primary endpoint (n=10) |
|---|---|---|
| TB Unit: Shirur / Bhor / Haveli PMC / Haveli PCMC | 2 / 0 / 9 / 4 | 1 / 0 / 7 / 2 |
| Age in years: median (range) | 34 (14 to 70) | 35.5 (17 to 59) |
| Aged under 18 years | 2 | 1 |
| Male / female | 9 / 6 | 6 / 4 |
| Site: pulmonary / extrapulmonary (pleural 3, abdominal 1) | 11 / 4 | 10 / 0 |
| Basis of diagnosis: Truenat / CBNAAT / chest radiograph / cytopathology | 7 / 3 / 4 / 1 | 7 / 3 / 0 / 0 |
| Recorded in Nikshay as bacteriologically confirmed | 12 | 10 |
| Key population recorded: urban slum / migrant / other or none | 10 / 1 / 4 | 7 / 0 / 3 |
| Notified by public / private sector | 14 / 1 | 10 / 0 |
| HIV non-reactive / not recorded | 14 / 1 | 9 / 1 |
| Treatment started within 3 days of diagnosis | 15 | 10 |
| Month of diagnosis: April / May 2026 | 2 / 13 | 1 / 9 |

Source: ACF case line list, final extract of 29 June 2026 (Nikshay episode records of all 15 cases). Two cases recorded as bacteriologically confirmed in Nikshay did not meet the primary definition: one pulmonary case with chest radiography as the recorded basis of diagnosis and no microbiological result, confirmed by the field team as clinically diagnosed (the Nikshay flag is an entry inconsistency), and one abdominal case diagnosed on cytopathology. Had the Nikshay flag been taken at face value, the comparison would have been 11 versus 9 (rate ratio 3.28, 95% CI 1.24 to 8.95; p=0.010); the primary count is 10. Rifampicin resistance was not recorded for any case. Nikshay residence fields differed from the sweep unit for several cases and were not used for attribution.

### S3. TREND checklist

| Item | TREND requirement | Where addressed |
|---|---|---|
| 1 | Title and abstract: information on how units were allocated; structured abstract; target population | Title; Summary (Design); Methods, Design and setting |
| 2 | Introduction: scientific background and explanation of rationale; theories used in designing the intervention | Introduction |
| 3 | Participants: eligibility criteria; recruitment method; sampling method; settings; recruitment and follow-up dates | Methods, Design and setting; AI-guided arm; Standard arm; S1.2 |
| 4 | Interventions: details of the intervention and comparison conditions | Methods, The planning tool; AI-guided arm; Standard arm; S1.1 |
| 5 | Objectives: specific objectives and hypotheses | Introduction, final paragraph |
| 6 | Outcomes: primary and secondary outcomes; methods used to collect data and enhance quality | Methods, Outcomes; S2.1 |
| 7 | Sample size: how sample size was determined | Methods, Sample size; S2.6 |
| 8 | Assignment method: unit of assignment; method of assignment; restrictions | Methods, Design and setting (planning strategies compared in the same units, non-randomised) |
| 9 | Blinding: whether participants, those administering the intervention or assessors were blinded | Not possible; outcome ascertainment used programme records (Methods, Outcomes) |
| 10 | Unit of analysis: description and how it relates to the unit of assignment | Methods, Sample size and statistical analysis (unit-level rates; pooled estimand) |
| 11 | Statistical methods: methods for primary outcomes, additional analyses, subgroups, missing data | Methods, Sample size and statistical analysis; S1.3 |

| Item | TREND requirement | Where addressed |
|---|---|---|
| 12 | Participant flow: numbers at each stage | Results, Screening cascade; Table 1; S2.1 |
| 13 | Recruitment: dates defining recruitment and follow-up | Methods, AI-guided arm (April to May 2026); Standard arm |
| 14 | Baseline data: characteristics of each condition | Methods, Design and setting; Table S1; S2.1 |
| 15 | Baseline equivalence: data on comparability of groups | Discussion, limitations (non-concurrent comparator, denominators) |
| 16 | Numbers analysed: number in each group included in each analysis; intention-to-treat | Table 2 (as-screened population) |
| 17 | Outcomes and estimation: summary of results with estimated effect size and precision | Results; Tables 1 and 2; Figure 1 |
| 18 | Ancillary analyses: subgroup and sensitivity analyses | Results (rural and urban; leave-one-out); S2.2 to S2.4 |
| 19 | Adverse events: important adverse events or unintended effects | Results, Acceptability (quota display, navigation failure, workload) |
| 20 | Interpretation: considering hypotheses, sources of bias, imprecision | Discussion, paragraphs 1 to 2 and limitations |
| 21 | Generalisability: external validity | Discussion, urban finding and limitations (single district) |
| 22 | Overall evidence: interpretation in the context of current evidence and theory | Discussion, paragraph 4 and final paragraph |

### S4. COREQ checklist

| Domain | Item | Where addressed |
|---|---|---|
| 1. Research team and reflexivity | 1 Interviewer or facilitator | A moderator and a note-taker from the implementation partner's field team at each session; names recorded on the FGD template (S1.5) |
| | 2 Credentials | Public health and field programme staff of the implementation partner |
| | 3 Occupation | FIND field coordinators and data analysts (Declarations, Contributors) |
| | 4 Gender | Not reported |

| Domain | Item | Where addressed |
|---|---|---|
| | 5 Experience and training | Not reported |
| | 6 Relationship established | Facilitators were known to participants through implementation; stated in Discussion, limitations |
| | 7 Participant knowledge of the interviewer | Participants knew the facilitators as the implementation team |
| | 8 Interviewer characteristics | Implementation-partner staff known to participants; the implementer's interest in the tool is stated as a limitation (Discussion) |
| 2. Study design | 9 Methodological orientation and theory | Structured descriptive approach: group-level ratings and feature votes, content synthesis of note-taker observations (S1.5) |
| | 10 Sampling | Purposive, all four units, workers who had received or used microplans (S1.5) |
| | 11 Method of approach | Invited through the TB Units |
| | 12 Sample size | 100 participants in 16 groups (Results; S2.5) |
| | 13 Non-participation | Not recorded |
| | 14 Setting of data collection | Not reported |
| | 15 Presence of non-participants | Not recorded |
| | 16 Description of sample | S2.5 |
| | 17 Interview guide | Structured five-part template with a hands-on demonstration (S1.5); available on request |
| | 18 Repeat interviews | None |
| | 19 Audio or visual recording | Responses recorded in writing by a designated note-taker on the structured template; sessions were not transcribed |
| | 20 Field notes | Note-taker observations and investigator field notes (FGD analysis report, version 3.0) |
| | 21 Duration | Not reported |
| | 22 Data saturation | Not assessed |
| | 23 Transcripts returned | Not applicable; no transcripts were produced |
| 3. Analysis | 24 Number of data coders | Two (MD, with HV) |

| Domain | Item | Where addressed |
|---|---|---|
| and findings | | |
| | 25 Description of the coding tree | Categories of the structured template; no inductive coding tree |
| | 26 Derivation of themes | Structured by the template's domains; note-taker observations synthesised by content (S1.5) |
| | 27 Software | Microsoft Excel (FGD data analysis tool, 18 May 2026) |
| | 28 Participant checking | Not performed |
| | 29 Quotations presented | One verbatim quotation in S2.5; responses were otherwise recorded as notes, not verbatim |
| | 30 Data and findings consistent | Results, Acceptability; Table 4 |
| | 31 Clarity of major themes | Results, Acceptability (efficiency, navigation, workload, quota display) |
| | 32 Clarity of minor themes | Results, Acceptability (Marathi outputs, plan-access overhead) |
